# Developing and validating a pancreatic cancer risk model for the general population using multi-institutional electronic health records from a federated network

**DOI:** 10.1101/2023.02.05.23285192

**Authors:** Kai Jia, Steven Kundrot, Matvey Palchuk, Jeff Warnick, Kathryn Haapala, Irving Kaplan, Martin Rinard, Limor Appelbaum

## Abstract

**Purpose:** Pancreatic Duct Adenocarcinoma (PDAC) screening can enable detection of early-stage disease and long-term survival. Current guidelines are based on inherited predisposition; only about 10% of PDAC cases meet screening eligibility criteria. Electronic Health Record (EHR) risk models for the general population hold out the promise of identifying a high-risk cohort to expand the currently screened population. Using EHR data from a multi-institutional federated network, we developed and validated a PDAC risk prediction model for the general US population.

**Methods:** We developed Neural Network (NN) and Logistic Regression (LR) models on structured, routinely collected EHR data from 55 US Health Care Organizations (HCOs). Our models used sex, age, frequency of clinical encounters, diagnoses, lab tests, and medications, to predict PDAC risk 6-18 months before diagnosis. Model performance was assessed using Receiver Operating Characteristic (ROC) curves and calibration plots. Models were externally validated using location, race, and temporal validation, with performance assessed using Area Under the Curve (AUC). We further simulated model deployment, evaluating sensitivity, specificity, Positive Predictive Value (PPV) and Standardized Incidence Ratio (SIR). We calculated SIR based on the SEER data of the general population with matched demographics.

**Results:** The final dataset included 63,884 PDAC cases and 3,604,863 controls between the ages 40 and 97.4 years. Our best performing NN model obtained an AUC of 0.829 (95% CI: 0.821 to 0.837) on the test set. Calibration plots showed good agreement between predicted and observed risks. Race-based external validation (trained on four races, tested on the fifth) AUCs of NN were 0.836 (95% CI: 0.797 to 0.874), 0.838 (95% CI: 0.821 to 0.855), 0.824 (95% CI: 0.819 to 0.830), 0.842 (95% CI: 0.750 to 0.934), and 0.774 (95% CI: 0.771 to 0.777) for AIAN, Asian, Black, NHPI, and White, respectively. Location-based external validation (trained on three locations, tested on the fourth) AUCs of NN were 0.751 (95% CI: 0.746 to 0.757), 0.749 (95% CI: 0.745 to 0.753), 0.752 (95% CI: 0.748 to 0.756), and 0.722 (95% CI: 0.713 to 0.732) for Midwest, Northeast, South, and West, respectively. Average temporal external validation (trained on data prior to certain dates, tested on data after a date) AUC of NN was 0.784 (95% CI: 0.763 to 0.805). Simulated deployment on the test set, with a mean follow up of 2.00 (SD 0.39) years, demonstrated an SIR range between 2.42-83.5 for NN, depending on the chosen risk threshold. At an SIR of 5.44, which exceeds the current threshold for inclusion into PDAC screening programs, NN sensitivity was 35.5% (specificity 95.6%), which is 3.5 times the sensitivity of those currently being screened with an inherited predisposition to PDAC. At a chosen high-risk threshold with a lower SIR, specificity was about 85%, and both models exhibited sensitivities above 50%.

**Conclusions:** Our models demonstrate good accuracy and generalizability across populations from diverse geographic locations, races, and over time. At comparable risk levels these models can predict up to three times as many PDAC cases as current screening guidelines. These models can therefore be used to identify high-risk individuals, overlooked by current guidelines, who may benefit from PDAC screening or inclusion in an enriched group for further testing such as biomarker testing. Our integration with the federated network provided access to data from a large, geographically and racially diverse patient population as well as a pathway to future clinical deployment.

## 1 Introduction

Most cases of Pancreatic Duct Adenocarcinoma (PDAC) are diagnosed as advanced-stage disease, leading to a five-year relative survival rate of only 11% [26]. Expanding the population currently being screened for this lethal disease is crucial for increasing early detection and improving survival. Current screening guidelines [4, 10, 12] targeting stage I cancers and high-grade PDAC precursors have been shown to significantly improve long-term survival [6, 18]. Current guidelines target patients with a family history or genetic predisposition to PDAC [13, 21], with screening eligibility based on estimated absolute and relative risk compared to the general population (5% or 5 times the relative risk, respectively) [6]. These patients comprise only about 10% of all PDAC cases. No consensus or guidelines exist for PDAC screening in the *general population* [20], where the *majority* of PDAC cases are found.

Several groups have developed PDAC risk models for the general population using various data sources [5, 15, 16]. A goal of most such models is eventual integration with Electronic Health Record (EHR) systems and ultimately clinical implementation. EHR integration has proven to be a significant barrier to the clinical adoption of models [28]. One effort developed a model using EHR data from an aggregated multi-institutional database [7]. The evaluation focused on identification of high risk patients up to one month before diagnosis and did not attempt to evaluate model generalization across locations or races. Several other efforts worked with real-world EHR data [3, 8, 22], but with limited validation across diverse locations and races. Other efforts worked with small sample sizes [5, 19] and internal validation only [16, 19].

We used EHR data from 55 US Health Care Organizations (HCOs) from a federated data network to develop and validate two PDAC risk prediction models for the general population, a Neural Network (NN) model and a Logistic regression (LR) model. The models can be used as a tool to identify individuals at high risk for PDAC from the general population, so they can be offered early screening or referred for lower overhead testing such as biomarker testing.

The network provides access to harmonized, de-identified EHR data of over 89 million patients for model development and testing. It also provides a means to simulate deployment of the resultant models to identify high risk patients for screening within a research setting. Because the network is connected to the EHR systems of the participating HCOs, it provides a pathway to deploy the models to a clinical setting, a critical step in the progression towards successful clinical adoption [28].

We developed a methodology to train PDAC prediction models on federated network EHR data. Our evaluation reports AUC and PPV numbers for the resulting trained models, with the evaluation focusing on the ability of the models to identify high risk patients 6 to 18 months before an initial PDAC diagnosis. We conducted three types of external validation: location-based, race-based, and temporal. We simulated deployment of the model on real-world HCO data to evaluate its performance in a more realistic setting. We compared the relative incidence of PDAC in our model-assigned high-risk group with that of a demographically matched general US population based on SEER data [1].

## 2 Methods

### 2.1 Data source and setting

This is an observational retrospective study, with both a case-control and cohort design, using data from the federated EHR database platform of TriNetX [27]. TriNetX is a federated global health research network that specializes in data collection and distribution. HCOs contributing to the database include academic medical centers, community hospitals, and outpatient clinics.

We used retrospective de-identified EHR data from 55 HCOs across the United States. The majority of these HCOs are tertiary care centers and the data used includes inpatient, outpatient, and Emergency Room encounters. Different HCOs have different historical coverage; on average, each HCO provides approximately 13 years of historical data. Data include values from structured EHR fields (e.g. demographics, date-indexed encounters, diagnoses, procedures, labs, and medications) as well as facts and narratives from free text (e.g. medications identified through Natural Language Processing (NLP)). TriNetX harmonizes all data from each HCO ‘s EHR to the TriNetX common data model and common set of controlled terminologies.TriNetX also has tools to identify anomalies and outliers for quality assurance.

We used data from the TriNetX database under a no-cost collaboration agreement between BIDMC, MIT, and TriNetX. Under this agreement, we accessed de-identified data under the agreements and institutional approvals already in place between TriNetX and their partner institutions.

### 2.2 Study population

We worked with two cohorts: a PDAC cohort and a control cohort. We obtained all data from TriNetX during November and December, 2022. We obtained the PDAC cohort by querying the TriNetX database to obtain EHR data for all patients, 40 years of age or older, from 55 HCOs across the United States, with one of the following ICD-10/ICD-9 codes:

- C25.0 Malignant neoplasm of head of pancreas
- C25.1 Malignant neoplasm of body of pancreas
- C25.2 Malignant neoplasm of tail of pancreas
- C25.3 Malignant neoplasm of pancreatic duct
- C25.7 Malignant neoplasm of other parts of pancreas
- C25.8 Malignant neoplasm of overlapping sites of pancreas
- C25.9 Malignant neoplasm of pancreas, unspecified
- 157 Malignant neoplasm of pancreas (ICD-9 without a corresponding ICD-10 code)

We obtained n=132,789 PDAC cases. We excluded patients who were diagnosed before 40 years of age (n=1,924), patients with no medical history 6 months prior to diagnosis (n=66,731), and patients with records 2 months after their death record (n=250), to obtain a PDAC cohort with n=63,884 cases.

To prepare the control cohort, we queried the TriNetX database for patients at least 40 years of age without any of the above ICD-10 or ICD-9 codes. There were n=51,139,587 patients that met this criteria. From these patients we randomly selected n=6,499,996 patients. We excluded patients with a PDAC tumor registry entry but no PDAC diagnosis entries (n=304), patients whose last entry was before age 35.5 (n=118,170), patients with less than 90 days of medical history (n=2,753,897), and patients with records 2 months after their death record (n=22,762), to obtain a control cohort with n=3,604,863 cases. Our subsequent training and testing procedures implement additional exclusion criteria (see below).

### 2.3 Model development

We used the TRIPOD guidelines for multivariable prediction models for reporting on model development and validation [9].

We trained and evaluated two model classes, Neural Network (NN) and Logistic Regression (LR). Data was randomly partitioned into training, validation, and test sets (75%, 10%, and 15%, respectively). We evaluated model performance by AUC scores and sensitivity, specificity, PPV, and SIR in simulated deployment. To calculate SIR, we used the SEER database [1] to estimate the PDAC risk for our model ‘s high-risk group compared to the general population.

Our training and testing procedures work with a cutoff date *C* for every patient, with entries after the cutoff date excluded. For a patient *P* and a cutoff date *C*, the model uses entries available before the cutoff date *C* to predict the risk of first diagnosis of PDAC between *C* + 6mo to *C* + 18mo. We defined the date of PDAC diagnosis *D* to be the first time a PDAC ICD code (as above) appeared in the patient record. During training, we sampled the cutoff dates for PDAC cases uniformly between [*D* - 18mo, *D* - 6mo]. Since control patients were not diagnosed with PDAC, we sampled random cutoff dates for them from the distribution of the PDAC diagnosis dates. For a control patient with a known death date, we limited the cutoff date to at most 18 months before death, to rule out undiagnosed PDAC that caused death. To avoid undiagnosed PDAC cases, we also limited all cutoff dates of patients in the control cohort to be at most 18 months before the dataset query date.

We empirically defined any patient with at least 16 diagnosis, medication, or lab result entries within 2 years before their cutoff date and whose first entry is at least 3 months earlier than their last entry before the cutoff date to have *sufficient medical history*. We excluded patients that did not have sufficient medical history. We trained the NN with the iterative Stochastic Gradient Descent (SGD) algorithm [17], sampling a new cutoff date for each patient at each step of the iteration. Our LR training sampled one cutoff date for each patient.

Our feature extraction excluded entries after the cutoff date (and included all entries up to the cutoff date). For each patient, we defined six basic features including age, whether age is known, sex, whether sex is known, number of diagnosis, medication, or lab entries in the medical record up to 18 months before cutoff (the recent entries), and number of diagnosis, medication, or lab entries in the medical record greater than 18 months before cutoff (the early entries). We also included features that correspond to individual diagnosis, medication, or lab codes, with the corresponding code empirically included in feature selection if it appeared in the medical record of at least 1% of the patients in the cancer cohort of the training set.

We manually grouped 827 commonly used diagnosis codes into 39 groups. For ungrouped codes, we used the ICD-10 category plus the first digit of the subcategory. We derived 3 features for each diagnosis code: whether or not it exists {0, 1}, its first and last date (encoding for first and last date: greater or equal to 4 years before cutoff=0; at cutoff=1). To use past ICD-9 data to train the model for use on current and future ICD-10 data, we mapped all ICD9 codes to their ICD-10 equivalents. For ICD-9 codes that could be mapped to more than one ICD-10 code, we included the features of all the mapped ICD-10 codes in the feature vector. We also manually grouped 67 medication codes into 8 different medication classes. Ungrouped codes were used as they are. We derived 4 features for each medication code: whether or not it exists {0, 1}, its frequency (i.e., number of times it appears in the medical record), span (time between first and last appearance of a medication code), and last date (same encoding as diagnosis first/last date). For lab features, we used a grouping provided by TriNetX for similar lab tests, which had 98 groups for 462 codes. Ungrouped codes were used as they are. For each lab code, we derived 4 features: existence, frequency, first date, and last date. The frequency was the number of lab results within three years before cutoff. We manually selected the most relevant lab tests for PDAC prediction, based on clinical knowledge and literature review. For these manually selected 44 quantitative labs, we derived two additional features: lab test value and slope. Lab values were normalized according to the median absolute deviation and the population median (range -1 to 1). Slope was measured by calculating the yearly change in lab test values up to three years before cutoff.

To account for the additional effect of the healthcare process on EHR data [2], we did the following: For each feature type described above (except the number of early and recent entries in basic features) there is a corresponding existence feature {0, 1} ; if the feature is missing in the data set, the value of the corresponding existence feature is 1 and the value of the feature itself is 0. This encoding enables the model to compute risk scores based on whether a feature is present or missing. Because our NN models can use sophisticated nonlinear reasoning to extract information from the chosen features, data imputation provides little to no useful additional information for these models. Therefore, we did not use any imputation.

Our NN models have three fully connected layers; each layer has 48, 16, and 1 output neurons. Hidden layers use the tanh nonlinearity. To ameliorate overfitting, we used sparse weights computed by the recently developed BinMask sparsification technique [14]. We used balanced numbers of PDAC and control patients in each mini-batch. For LR training, we used the SAGA solver [11] with balanced class weights. For each model type, we trained four models with different regularization parameters and selected the best one on the validation set.

We calibrated the models on the validation set with a modified Platt calibration algorithm [23], where we fitted a two-segment piecewise-linear mapping with the turning point set as the median of model predictions. We accounted for the unbalanced sampling of control cohort and estimated the risk on the whole population in calibration. We evaluated our calibration by creating calibration plots on the test set. We chose 16 risk groups for calibration evaluation as a geometric sequence between the 85% percentile of predicted risk on the test set and the maximum predicted risk. To quantitatively compare calibration between models, we used the Geometric Mean of Over Estimation (GMOE), calculated as the geometric mean of the ratios of predicted risk to the true risk over all tested risk groups. Perfectly calibrated models have GMOE=1. A GMOE value greater than one means over estimation of risk and a value less than one means under estimation of risk.

We also evaluated the stability of our algorithm by calculating the mean AUC and GMOE with confidence interval on nine independent runs with different random seeds for dataset split and weight initialization. For both the LR and NN models, we analyzed the impact of different numbers of features on model performance. We reduced the number of input features by applying BinMask to the input of a small and densely connected neural network to automatically select important features. We varied the BinMask weight decay coefficient to obtain different numbers of input features and evaluated the performance of our models with those feature sets.

We analyzed the feature importance for NN by calculating the partial AUC (up to 6% FPR) obtained with only each type of medical record entries. A larger score for a type of record means the NN makes better predictions based on the record entries alone.

### 2.4 External validation

Our model validation considered three attributes: geographical location of the HCO, patient race, and time of diagnosis/last used entry in the medical record. For each attribute, we split the dataset according to that attribute, trained models on one split, and tested on the other split.

Our location based validation used the TriNetX geographical location for each HCO; locations include Northeast, South, Midwest, and West. Our race based validation used the TriNetX racial classification of each patient; races include American Indian or Alaska Native (AIAN), Asian, Black or African American (Black), Native Hawaiian or Other Pacific Islander (NHPI), and White.

A primary assessment of model generalizability is the AUC gap between test set and validation set. However, since different attribute splitting produces training/validation/test sets with different sizes, the test/validation AUC gap does not necessarily depict model generalizability. Therefore, we trained control models that used the same training and test set size for each attribute-based split, but used random splitting that ignores attribute values. We also assessed model generalizability by checking the AUC gap between the external validation models and corresponding control models.

For temporal validation, we selected the 50%, 60%, …, 90% percentile from the distribution of diagnosis dates as the dataset split dates. The 90% percentile was Sep 23, 2021. We trained the models only on data available prior to those split dates. We also limited the cutoff date of control patients to earlier than 18 months before the split dates, to simulate model training with datasets queried on the split dates. We evaluated the performance of the models on the same subset of data only available after Sep 23, 2021. We also calculated the average performance of different models for the temporal validation. Since different dataset split dates result in different training set sizes, we also trained control models. For each split date, we randomly sampled the same number of PDAC cases (equal to the 50% of the total number of PDAC cases) from cases up to that split date. The control models allowed us to separate the contribution of larger training set from the impact of smaller time gap between training and test sets.

### 2.5 Simulated deployment

We estimate the performance of our model when deployed in a clinical setting by simulating model deployment in a prospective study on the TriNetX database. We trained the model only on data available prior to Feb 7, 2020, in the same way as the above temporal validation, with the dataset split date chosen as the 70% percentile of the distribution of the diagnosis dates. For each date *D* separated by 90 days between Feb 7, 2020 and May 2, 2021 (18 months before dataset query), we

1. Enrolled a new patient into the simulated deployment if the patient had a known age, was at least 40 years old on date *D*, and had sufficient medical history on *D* for the first time. We call the date *D* the *enrollment date* for such a patient.
2. For each enrolled patient, we checked if that patient still had sufficient medical history on *D*. If so, we evaluated the model risk by our model, with the cutoff date set at *D*. We call the date *D* a *check date* for such a patient.

We excluded patients who were diagnosed with PDAC either before enrollment or within 6 months after enrollment, patients who had no medical entries between first and last check dates, and patients with a known death but no PDAC diagnosis within 18 months after enrollment. We started following up a patient 6 months after their enrollment date. We stopped following up a patient 18 months after the last check date. During the followup period, we defined the following outcomes:

1. A patient was diagnosed with PDAC. We counted this patient as a true positive if the model made a high-risk prediction on any check date 6 months prior to diagnosis and a false negative otherwise.
2. A patient was not diagnosed with PDAC. They might either have a known death date, reached our dataset query date, or never had sufficient medical history again after a certain check date. For patients with a known death date, we only considered check dates up to 18 months before death, due to the possibility of undiagnosed PDAC at death. For other patients, we considered all check dates. If the model ever made a high-risk prediction for this patient on any considered check dates, we counted the patient as a false positive. Otherwise, we counted the patient as a true negative.

We chose the risk thresholds according to the 89.00%, 93.00%, 96.50%, 98.00%, 99.70%, 99.92% specificity levels on the validation set. For each risk threshold, we computed sensitivity, specificity, Positive Predictive Value (PPV), and Standardized Incidence Ratio (SIR), based on the above protocol. Since we used all the PDAC cases in the TriNetX database, but sampled a subset of control patients, we accounted for this imbalance to estimate the PPV and SIR that would be obtained if we had evaluated the model on the full TriNetX population.

We calculated SIR by dividing the observed PDAC cases in the high-risk group by the expected number of PDAC cases of that group. To calculate the expected number of cases, we used the SEER database [1], matched with age, sex, race, and calendar year for each individual in the high-risk group, as done by Porter et al. [24].

## 3 Results

### 3.1 Model evaluation

The final LR model and NN models used 63,884 cancer patients and 3,604,863 controls up to 97.4 years old (determined at the time of diagnosis or last record). Detailed demographics, including sex, age, race, and HCO location, are given in Table 1. Fig. 1 presents a flowchart demonstrating how this dataset was derived.

**Table 1:** Demographics of our dataset.

| Attribute |  | Cancer group (n=63,884)<br>% (No.) | Control group (n=3,604,863)<br>% (No.) |
| --- | --- | --- | --- |
| Sex | Female | 50.40 (32,196) | 55.27 (1,992,432) |
|  | Male | 49.59 (31,681) | 43.42 (1,565,131) |
|  | Unknown | 0.01 (7) | 1.31 (47,300) |
| Age at first record | Mean (SD) | 60.88 (12.02) | 53.90 (14.03) |
|  | < 40 | 4.88 (3,116) | 17.37 (626,073) |
|  | 40 - 50 | 12.38 (7,908) | 21.52 (775,841) |
|  | 50 - 60 | 24.35 (15,556) | 23.30 (840,092) |
|  | 60 - 70 | 30.69 (19,605) | 19.76 (712,411) |
|  | 70 - 80 | 18.93 (12,091) | 11.01 (396,806) |
|  | > 80 | 3.54 (2,259) | 2.50 (90,128) |
| Age at diagnosis<br>/ last record | Mean (SD) | 67.67 (10.59) | 60.20 (13.10) |
|  | < 40 | 0.00 (0) | 4.78 (172,349) |
|  | 40 - 50 | 6.01 (3,841) | 20.00 (720,800) |
|  | 50 - 60 | 16.42 (10,490) | 22.90 (825,442) |
|  | 60 - 70 | 30.56 (19,522) | 23.44 (844,818) |
|  | 70 - 80 | 29.68 (18,958) | 17.05 (614,615) |
|  | > 80 | 12.09 (7,724) | 7.30 (263,327) |
| Age | Unknown | 5.24 (3,349) | 4.54 (163,512) |
| Race | AIAN | 0.26 (164) | 0.36 (13,023) |
|  | Asian | 1.53 (976) | 2.27 (81,726) |
|  | Black | 13.95 (8,910) | 14.16 (510,444) |
|  | NHPI | 0.05 (35) | 0.13 (4,694) |
|  | White | 72.70 (46,441) | 67.24 (2,423,771) |
|  | Unknown | 11.52 (7,358) | 15.85 (571,205) |
| HCO location | Midwest | 21.17 (13,527) | 15.41 (555,417) |
|  | Northeast | 33.42 (21,352) | 28.40 (1,023,916) |
|  | South | 36.37 (23,234) | 44.18 (1,592,634) |
|  | West | 7.41 (4,733) | 8.54 (308,013) |
|  | Unknown | 1.62 (1,038) | 3.46 (124,883) |
| No. medical records | Mean (SD) | 779.11 (1506.23) | 441.79 (1091.31) |

Race abbreviations:
- AIAN: American Indian or Alaska Native - Black: Black or African American - NHPI: Native Hawaiian or Other Pacific Islander

**Fig. 1:**
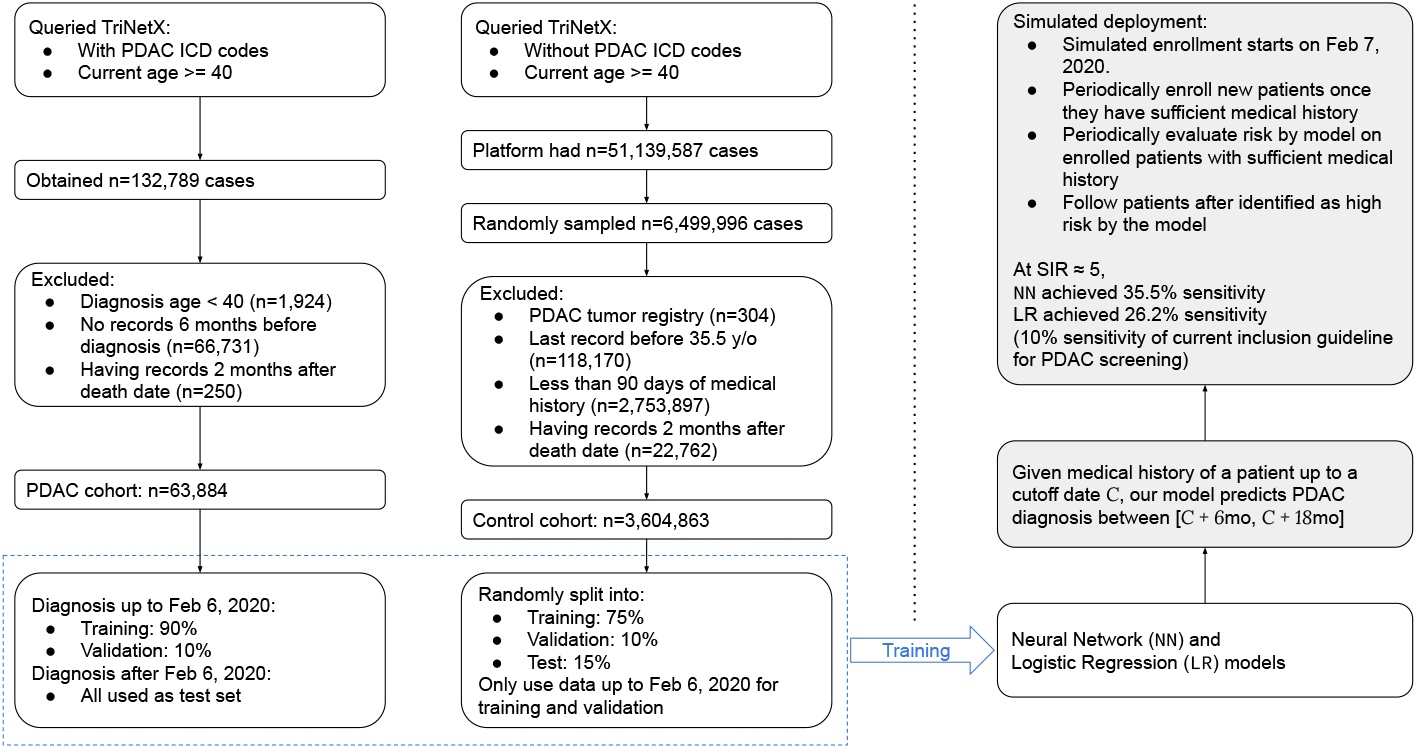
Flowchart of our study with simulated deployment as an example

The NN outperformed the LR model on the test set, with an AUC of 0.827 (95% CI: 0.822 to 0.833) and 0.809 (95% CI: 0.804 to 0.815), respectively (Fig. 2a). The mean AUCs of NN and LR on nine random runs are 0.829 (95% CI: 0.821 to 0.837) and 0.810 (95% CI: 0.803 to 0.817), respectively. Because our models predict based in part on the presence or absence of features, each feature is a predictor and we have no participants with missing predictors [2].

**Fig. 2:**
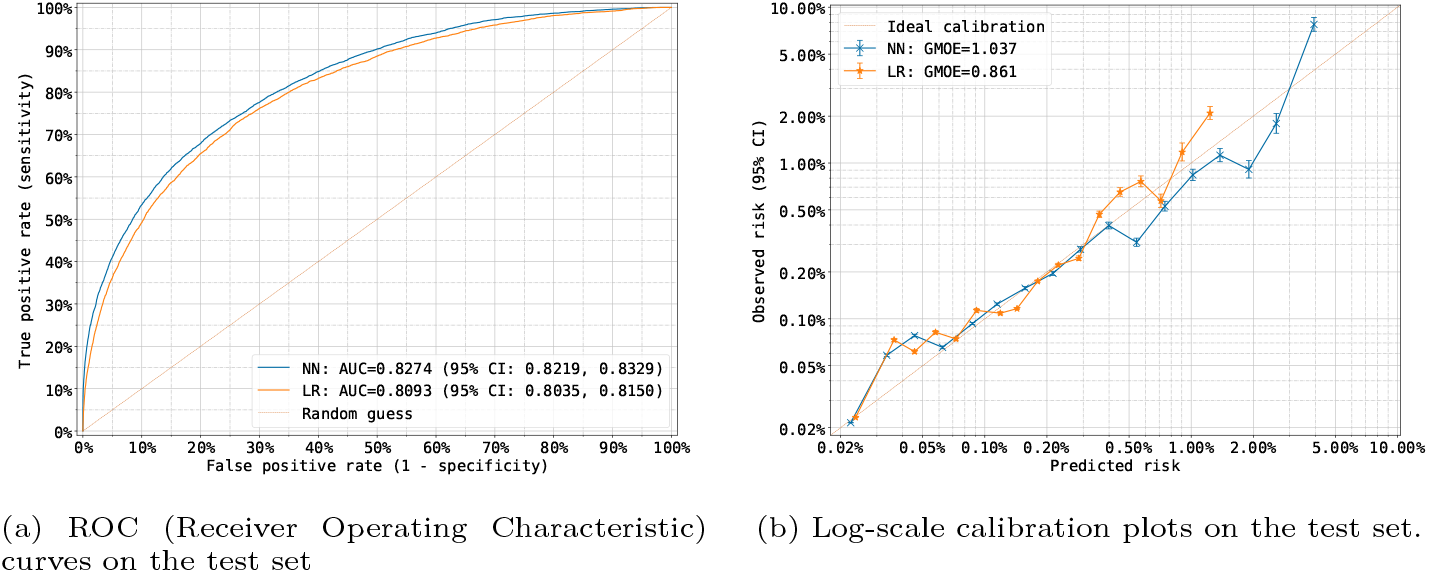
Model performance on the test set.

Fig. 2b shows the log-scale calibration plots on the test set. The evaluated risk levels are selected according to a geometric sequence between the 85% risk percentile and the maximal risk given by the model on the test set. Geometric Mean of Over Estimation (GMOE), the geometric mean of ratios of predicted risks to observed risks, was calculated for both models. The GMOE for the NN was 1.037 and 0.861 for the LR. The GMOE on nine random runs was 1.148 (95% CI: 1.092 to 1.203) and 0.992 (95% CI: 0.944 to 1.041) for NN and LR, respectively.

The impact of different feature numbers on model performance, for both the NN and LR models, is shown in Fig. 3a. Both models showed improved performance with an increasing number of features, reaching a plateau at an AUC of 0.83 (NN) and 0.81 (LR) for a combination of 1574 diagnoses features, 862 medication features, and 719 lab features. Additional features produced no significant improvement in model performance.

**Fig. 3:**
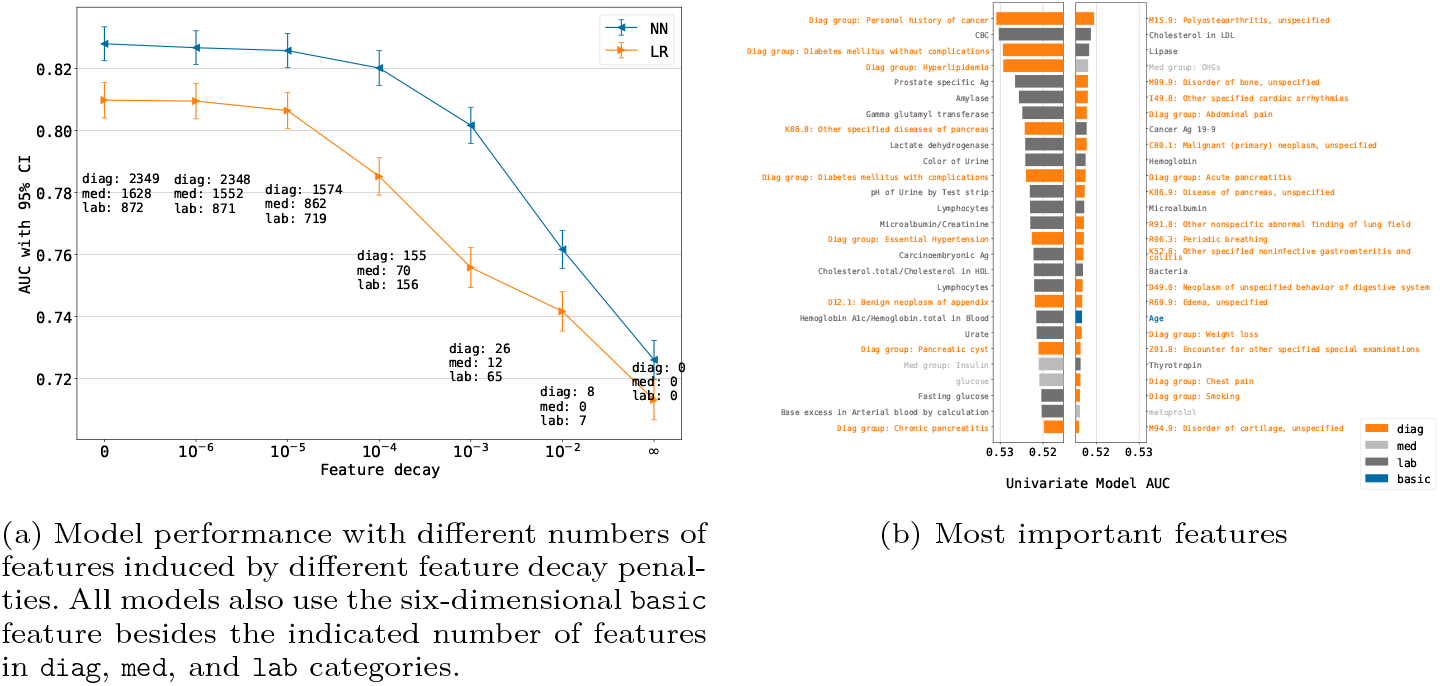
Feature analysis. In the plots, diag refers to diagnosis features, med refers to medication features, and lab refers to lab features.

Fig. 3b shows the top features selected by the LR model and ranked by feature importance. The top features include codes related to glucose metabolism and diabetes, medications such as Insulin and oral hypoglycemics, as well as blood tests for glucose and fasting glucose and HbA1c. Top features also include known PDAC risk factors such as age, pancreatitis, pancreatic cysts, personal history of cancer, weight loss, and smoking.

### 3.2 External validation results

Fig. 4 shows the results for race-based, location-based, and temporal external validations. The model performed similarly across racial groups without significant performance drop, as shown in Fig. 4a. AUCs on the test set were 0.836 (95% CI: 0.797 to 0.874), 0.838 (95% CI: 0.821 to 0.855), 0.824 (95% CI: 0.819 to 0.830), 0.842 (95% CI: 0.750 to 0.934), and 0.774 (95% CI: 0.771 to 0.777) for AIAN, Asian, Black, NHPI, and White racial groups, respectively. The AUCs of the LR models were 0.801 (95% CI: 0.755 to 0.846), 0.822 (95% CI: 0.804 to 0.840), 0.806 (95% CI: 0.800 to 0.811), 0.836 (95% CI: 0.742 to 0.929), and 0.773 (95% CI: 0.770 to 0.775). Test AUCs of NN models were -0.035 to 0.015 lower than the corresponding control models, and -0.024 to 0.008 lower for LR models. The number of patients of each racial groups can be seen in Table 1. We excluded patients with unknown race from this experiment.

**Fig. 4:**
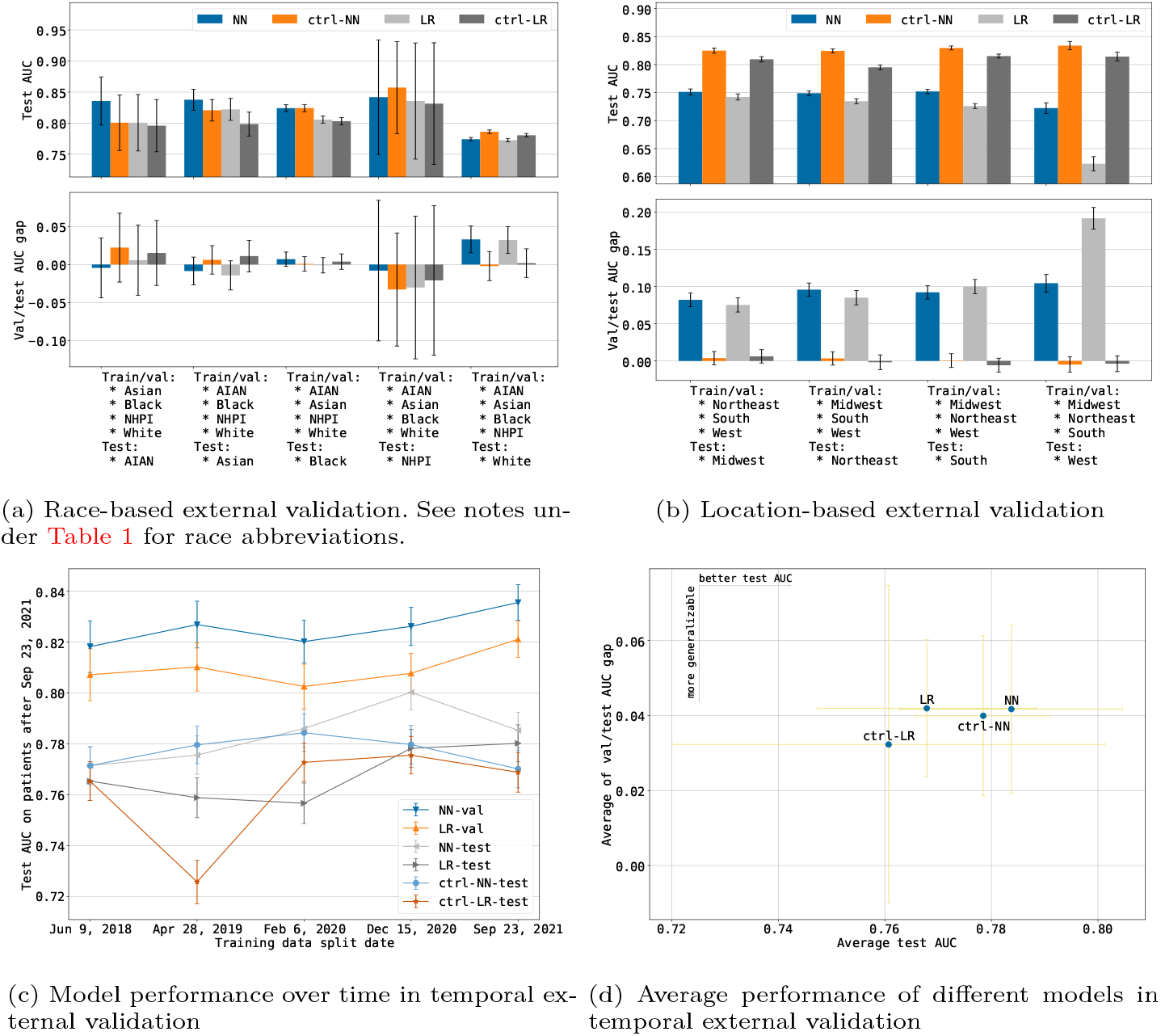
Results for location-based, race-based, and temporal external validations. Error bars indicate 95% CI.

Model performance was similar across the different geographic locations as shown in Fig. 4b. NN AUCs on the test set were 0.751 (95% CI: 0.746 to 0.757), 0.749 (95% CI: 0.745 to 0.753), 0.752 (95% CI: 0.748 to 0.756), and 0.722 (95% CI: 0.713 to 0.732) for the Midwest, Northeast, South, and West, respectively. LR AUCs were 0.742 (95% CI: 0.737 to 0.748), 0.735 (95% CI: 0.730 to 0.739), 0.726 (95% CI: 0.722 to 0.730), and 0.623 (95% CI: 0.610 to 0.636). Test AUCs of NN models were 0.074 to 0.112 lower than the corresponding control models, and 0.060 to 0.191 lower for LR models. The number of patients in each location can be seen in Table 1. We excluded patients with unknown HCO location from this experiment.

For temporal validation, model test performance varied over time, although they had relatively stable validation AUCs. Both NN and LR showed improved performance by adding more recent training data. The control models had worse performance and showed less stable improvement over time, which suggests that training set size is an important factor. The average test AUCs were 0.784 (95% CI: 0.763 to 0.805) and 0.768 (95% CI: 0.747 to 0.788) for the NN and LR models, respectively.

### 3.3 Simulated deployment results

The simulated deployment of the NN and LR models was on 201,703 patients (with 8,113 PDAC cases) in the test set, with enrollment from Feb 7, 2020 to May 2, 2021. Mean age at enrollment was 61.45 (SD 11.97). Mean age at PDAC diagnosis was 69.65 (SD 10.40). Each patient was followed up for 2.00 (SD 0.39) years (Table 2).

**Table 2:**
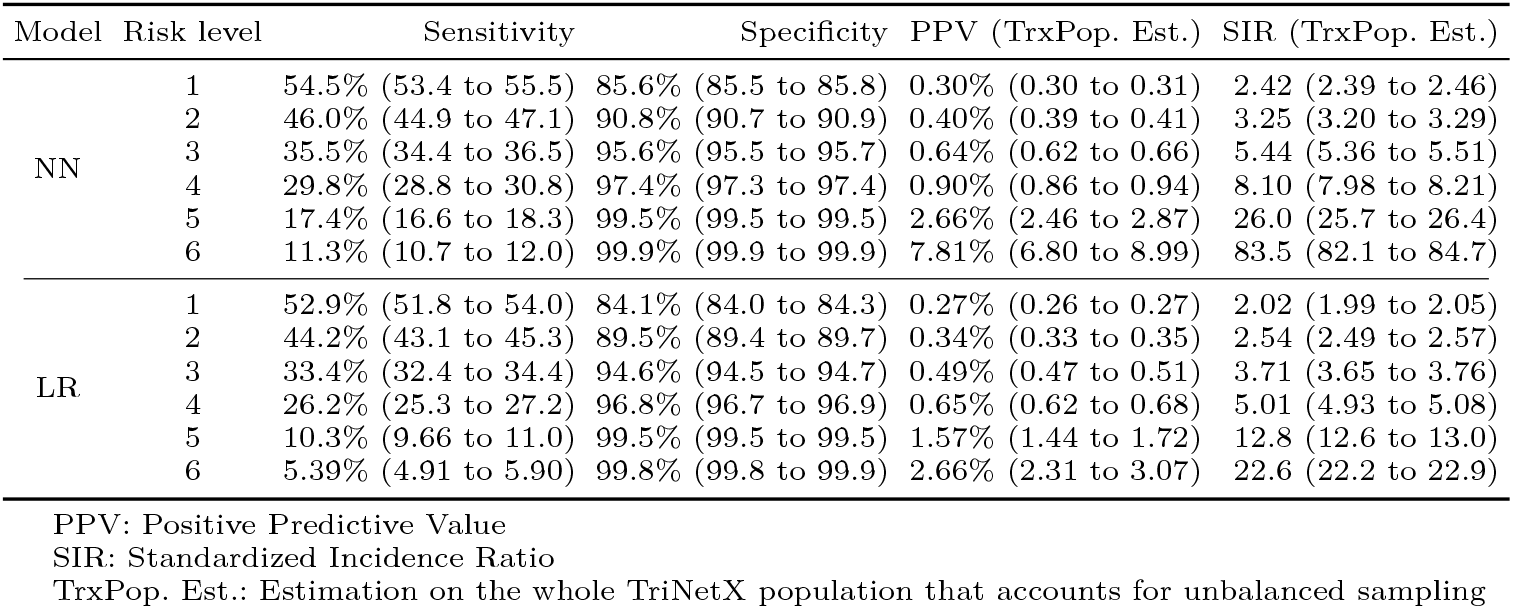
Simulated deployment results. Numbers in brackets are 95% CI.

Having accounted for unbalanced sampling of PDAC and control cohorts, we estimated that the model PPV range on the whole TriNetX population was 0.30%-7.81% for the NN and 0.27%-2.66% for the LR. NN and LR SIR ranges were 2.42-83.5 and 2.02-22.6, respectively. The SIR of all the enrolled patients during the follow-up period was 0.95 (95% CI: 0.94 to 0.96). An SIR close to 1 indicates that our TriNetX test population with patient exclusion has similar PDAC incidence as the general US population.

We determined the high-risk group to be any individuals that have an SIR of 5.44 or above, based on the NN model. This threshold is correlated with a 35.5% sensitivity and 95.6% specificity. We use this SIR threshold because it is similar to the currently used eligibility cutoff for inclusion of individuals into screening programs [12].

## 4 Discussion

Our study leveraged routinely collected EHR data from a federated network including 55 HCOs across the United States to develop and validate two ML models (NN and LR) that can accurately identify patients in the general population at high risk for PDAC, 6 to 18 months before first PDAC diagnosis. Both models were trained on 63,884 PDAC cases and 3,604,863 controls; both models worked with features derived from medical record entries including diagnosis, medication, and lab results, as well as basic features including sex, age, and number of clinical encounters. Our NN model obtained an AUC of 0.829 (95% CI: 0.821 to 0.837) on the test set; the LR model obtained an AUC of 0.810 (95% CI: 0.803 to 0.817).

### 4.1 Potential use cases

We anticipate two potential clinical use cases for our models. The first is to expand the eligibility for current screening programs, which are based on imaging modalities such Endoscopic UltraSound (EUS) and MRI/MRCP [6]. Current eligibility criteria are based on familial PDAC or a known germline mutation syndrome (e.g., Lynch, Peutz-Jeghers) [6]. The identified population is known to have an SIR of minimum 5 times the SIR of the general population and includes only 10% of PDAC cases [13, 21]. Depending on the chosen high-risk threshold, our NN model exhibited an SIR of 2.42 to 83.5. At an SIR of 5.44, our NN model identifies 35.5% of the PDAC cases as high risk 6 to 18 months before diagnosis, a significant improvement over current screening criteria.

The second use case is to identify an enriched group for lower overhead testing (such as biomarker testing) followed by screening based on the lower overhead test. In this use case we anticipate that it will be feasible to deploy the model at a higher sensitivity than in our first use case. For example, at 85.6% specificity, NN exhibited 54.5% sensitivity.

### 4.2 Race-based, location-based, and temporal validation

Our race based validation worked with the five racial groups recorded within the TriNetX EHR data: AIAN, Asian, Black, NHPI, and White. We trained models on four of these five racial groups, then tested on the fifth. The results showed similar performance across all training/test pairs, highlighting the generalizability across diverse racial populations. There was a small AUC drop for models when trained on all groups except White and tested on White, which we attribute to the fact that the White group included over 70% of the PDAC cases in the data set.

Our location based validation divided the HCOs into four regions: Midwest, Northeast, South, and West. We trained models on three of the regions, then tested on the fourth. In comparison with models trained on all regions with randomly sampled size-matched training data, these models showed modest AUC drops (0.074 to 0.112 for NN and 0.060 to 0.191 for LR).

Our temporal validation trained models on data before different dataset split dates, then tested the models on future dates not used for training. We found that NN models outperformed LR models, exhibiting average AUCs 0.784 (95% CI: 0.763 to 0.805) and 0.768 (95% CI: 0.747 to 0.788), respectively.

### 4.3 Simulated deployment

We envision the eventual deployment of our models into clinical practice to improve patient outcomes by promoting the detection of early stage disease. We evaluated the effectiveness of our models for this purpose by simulating the deployment of our models. A key aspect of this simulated deployment was using models trained only on data available before a simulated enrollment date to identify high-risk individuals after the simulated enrollment date. We then followed the identified high-risk individuals over time to evaluate the performance of our models.

This simulated deployment methodology stands in contrast to methodologies used in previous studies that do not temporally separate the training and test data [5, 7]. By more closely tracking the envisioned deployment scenario, we eliminated a potential source of inaccuracy and hope to obtain a more accurate prediction of model performance in clinical use.

### 4.4 Federated network

A significant strength of our work is the development and validation of our models using a federated EHR network. This network ingests EHR data from multiple HCOs and EHR sources, with the data remaining stored behind each institution ‘s firewall. The ingested data is de-identified, harmonized, and converted into a single format, supporting ease of integration and deployment of models within the same platform. This federated network enabled us to train and externally validate our models on racially, geographically, and temporally diverse data from 55 HCOs within the United States. The results show that our models perform well on all geographic and racial groups and generalize well across time. The network also enabled us to simulate deployment of the model over time to identify high-risk individuals across the entire network. The eventual clinical deployment of PDAC risk prediction models depends not only on model accuracy and generalizability, but also on productive integration into EHR systems for inclusion into the clinical workflow. Lack of system integration and model automation comprises a significant barrier to clinical adoption of such models [28]. Because of their close integration with existing HCO EHR systems, federated networks can solve these integration and deployment challenges to provide a clear pathway for integrated model development, validation, and clinical deployment all within a single federated system [25].

### 4.5 Related work

Other researchers have used EHR data to develop PDAC risk prediction models for the general population [3, 5, 7, 8, 22]. Data set sizes range from 1,792 PDAC cases/1.8M controls [8] to 24,000 PDAC cases/6.2M controls [22]. Some studies lack an external validation [7], complete the external validation/evaluate model generalizability only with data from a single geographic area [3, 22], or validate only on one gender (male) [8] or race [15]. While some studies work with data obtained from multiple organizations [7, 8, 22], none work with a federated data network that harmonizes and standardizes the data, none provides a clear path to clinical deployment, and none supports the seamless deployment of the model to new HCOs as they join the federated network.

Some previous studies evaluate the ability of their models to identify high-risk individuals either until or shortly before the date of PDAC diagnosis [7, 8, 22], when clinical benefit is improbable. To focus on time frames in which detection of early stage disease and potential cure are most likely, we evaluate the ability of our models to identify high-risk patients at least six months before diagnosis.

### 4.6 Limitations

Our study has limitations. Notably, model development and validation were retrospective. Prospective studies are needed to evaluate efficacy of clinical detection of early stage disease in high-risk individuals.

Our results also show that our models performed well on data from the TriNetX network, including multiple HCOs located in different geographic regions across the United States. We do not know, however, if our models will perform similarly on data from different sources or different countries. Future work should evaluate the models on data from different EHR sources and populations selected from different countries and global regions.

The use of neural networks and the fact that our model needed thousands of features to reach its best performance make it harder to interpret the reasoning process or extract knowledge for clinicians. Future work should try to gain a deeper understanding of how the model makes predictions and to simplify the model if possible.

## 5 Conclusion

In conclusion, we have built, validated, and simulated deployment of a PDAC risk prediction model for the general population on multi-institutional EHR data from a federated network. This model can be used to help primary care providers across the country identify high-risk individuals for PDAC screening or used as a first filter before subsequent biomarker testing. The model maintained its accuracy across diverse racial groups and geographic regions in the US, as well as over time, and outperformed widely-used clinical guideline criteria [10, 12] for inclusion of individuals into PDAC screening programs.

Our approach enables potential expansion of the population targeted for screening beyond the traditionally screened minority with an inherited predisposition. To our knowledge, this is the first PDAC risk prediction model developed, externally validated, with simulated deployment, using a federated network. The developed models set the stage for deployment of the model within the network to identify high risk patients at multiple institutions within the network. A prospective study to validate the models before full clinical deployment is the next step.

## Supporting information

TRIPOD Checklist: Prediction Model Development and Validation

## Data Availability

The de-identified data in TriNetX federated network database can only be accessed by researchers that are either part of the network or have a collaboration agreement with TriNetX. As stated in the manuscript, we accessed data as part of a no-cost collaboration agreement between BIDMC, MIT, and TriNetX.

## Acknowledgment

We are grateful to Gadi Lachman and TriNetX for providing support and resources for this work. We thank Lydia González for her help on identifying and mitigating data quality issues. We also thank the Prevent Cancer Foundation for supporting this work (LA).

## Author contributions

LA, MR, KJ, SK conceptualization. Data acquisition KH, JW, KJ. Data curation KJ, MR, LA. Data interpretation KJ, MR, LA, MP, IDK. Project administration LA, MR, KH. Supervision MR, LA, SK, MP. ALL writing review and editing. ALL approved published version and agreed to be accountable for all aspects of the work.

## References

1. Surveillance, epidemiology, and end results (SEER) program SEER*Stat database: Incidence SEER research limited-field data, 22 registries, nov 2021 sub (2000-2019) linked to county attributes time dependent (1990-2019) income/rurality, 1969-2020 counties (2022), https://www.seer.cancer.gov, Released April 2022, based on the November 2021 submission

2. Agniel, D., Kohane, I.S., Weber, G.M.: Biases in electronic health record data due to processes within the healthcare system: retrospective observational study. BMJ 361 (2018)

3. Appelbaum, L., Cambronero, J.P., Stevens, J.P., Horng, S., Pollick, K., Silva, G., Haneuse, S., Piatkowski, G., Benhaga, N., Duey, S., et al.: Development and validation of a pancreatic cancer risk model for the general population using electronic health records: An observational study. European Journal of Cancer 143, 19–30 (2021)

4. Aslanian, H.R., Lee, J.H., Canto, M.I.: Aga clinical practice update on pancreas cancer screening in high-risk individuals: expert review. Gastroenterology 159(1), 358–362 (2020)

5. Baecker, A., Kim, S., Risch, H.A., Nuckols, T.K., Wu, B.U., Hendifar, A.E., Pandol, S.J., Pisegna, J.R., Jeon, C.Y.: Do changes in health reveal the possibility of undiagnosed pancreatic cancer? development of a risk-prediction model based on healthcare claims data. PloS one 14(6), e0218580 (2019)

6. Canto, M.I., Harinck, F., Hruban, R.H., Offerhaus, G.J., Poley, J.W., Kamel, I., Nio, Y., Schulick, R.S., Bassi, C., Kluijt, I., et al.: International cancer of the pancreas screening (CAPS) consortium summit on the management of patients with increased risk for familial pancreatic cancer. Gut 62(3), 339–347 (2013)

7. Chen, Q., Cherry, D.R., Nalawade, V., Qiao, E.M., Kumar, A., Lowy, A.M., Simpson, D.R., Murphy, J.D.: Clinical data prediction model to identify patients with early-stage pancreatic cancer. JCO Clinical Cancer Informatics 5, 279–287 (2021)

8. Chen, W., Zhou, Y., Xie, F., Butler, R.K., Jeon, C.Y., Luong, T.Q., Lin, Y.C., Lustigova, E., Pisegna, J.R., Kim, S., et al.: Prediction model for detection of sporadic pancreatic cancer (pro-tect) in a population-based cohort using machine learning and further validation in a prospective study. medRxiv (2022)

9. Collins, G.S., Reitsma, J.B., Altman, D.G., Moons, K.G.: Transparent reporting of a multivariable prediction model for individual prognosis or diagnosis (tripod): the tripod statement. Journal of British Surgery 102(3), 148–158 (2015)

10. Daly, M.B., Pal, T., AlHilli, Z., Arun, B., Buys, S.S., Cheng, H., Churpek, J., Domchek, S.M., Elkhanany, A., Friedman, S., Giri, V., Goggins, M., Hagemann, A., Hendrix, A., Hutton, M.L., Karlan, B.Y., Kassem, N., Khan, S., Klein, C., Kohlmann, W., Kurian, A.W., Laronga, C., Mak, J.S., Mansour, J., Maxell, K., McDonnell, K., Menendez, C.S., Merajver, S.D., Norquist, B.S., Offit, K., Reiser, G., Senter-Jamieson, L., Shannon, K.M., Shatsky, R., Visvanathan, K., Welborn, J., Wick, M.J., Yurgelun, M.B., et al.: Genetic/familial high-risk assessment: Breast, ovarian, and pancreatic (2023), https://www.nccn.org/professionals/physician_gls/pdf/genetics_bop.pdf, Accessed: 1-21-2023

11. Defazio, A., Bach, F., Lacoste-Julien, S.: Saga: A fast incremental gradient method with support for non-strongly convex composite objectives. Advances in neural information processing systems 27 (2014)

12. Goggins, M., Overbeek, K.A., Brand, R., Syngal, S., Del Chiaro, M., Bartsch, D.K., Bassi, C., Carrato, A., Farrell, J., Fishman, E.K., et al.: Management of patients with increased risk for familial pancreatic cancer: updated recommendations from the international cancer of the pancreas screening (caps) consortium. Gut 69(1), 7–17 (2020)

13. Humphris, J.L., Johns, A.L., Simpson, S.H., Cowley, M.J., Pajic, M., Chang, D.K., Nagrial, A.M., Chin, V.T., Chantrill, L.A., Pinese, M., et al.: Clinical and pathologic features of familial pancreatic cancer. Cancer 120(23), 3669– 3675 (2014)

14. Jia, K., Rinard, M.: Efficient exact verification of binarized neural networks. In: Larochelle, H., Ranzato, M., Hadsell, R., Balcan, M.F., Lin, H. (eds.) Advances in Neural Information Processing Systems, vol. 33, pp. 1782–1795, Curran Associates, Inc. (2020)

15. Kim, J., Yuan, C., Babic, A., Bao, Y., Clish, C.B., Pollak, M.N., Amundadottir, L.T., Klein, A.P., Stolzenberg-Solomon, R.Z., Pandharipande, P.V., et al.: Genetic and circulating biomarker data improve risk prediction for pancreatic cancer in the general population. Cancer Epidemiology, Biomarkers & Prevention 29(5), 999–1008 (2020)

16. Klein, A.P., Lindström, S., Mendelsohn, J.B., Steplowski, E., Arslan, A.A., Bueno-de Mesquita, H.B., Fuchs, C.S., Gallinger, S., Gross, M., Helzlsouer, K., et al.: An absolute risk model to identify individuals at elevated risk for pancreatic cancer in the general population. PloS one 8(9), e72311 (2013)

17. LeCun, Y., Bengio, Y., Hinton, G.: Deep learning. nature 521(7553), 436–444 (2015)

18. Lu, C., Xu, C.F., Wan, X.Y., Zhu, H.T., Yu, C.H., Li, Y.M.: Screening for pancreatic cancer in familial high-risk individuals: A systematic review. World journal of gastroenterology: WJG 21(28), 8678 (2015)

19. Muhammad, W., Hart, G.R., Nartowt, B., Farrell, J.J., Johung, K., Liang, Y., Deng, J.: Pancreatic cancer prediction through an artificial neural network. Frontiers in Artificial Intelligence 2, 2 (2019)

20. Owens, D.K., Davidson, K.W., Krist, A.H., Barry, M.J., Cabana, M., Caughey, A.B., Curry, S.J., Doubeni, C.A., Epling, J.W., Kubik, M., et al.: Screening for pancreatic cancer: Us preventive services task force reaffirmation recommendation statement. Jama 322(5), 438–444 (2019)

21. Petersen, G.M.: Familial pancreatic cancer. In: Seminars in oncology, vol. 43, pp. 548–553, Elsevier (2016)

22. Placido, D., Yuan, B., Hjaltelin, J.X., Haue, A.D., Chmura, P.J., Yuan, C., Kim, J., Umeton, R., Antell, G., Chowdhury, A., Franz, A., Brais, L., Andrews, E., Marks, D.S., Regev, A., Kraft, P., Wolpin, B.M., Rosenthal, M., Brunak, S., Sander, C.: Pancreatic cancer risk predicted from disease trajectories using deep learning. BioRxiv (2021), https://doi.org/10.1101/2021.06.27.449937

23. Platt, J., et al.: Probabilistic outputs for support vector machines and comparisons to regularized likelihood methods. Advances in large margin classifiers 10(3), 61–74 (1999)

24. Porter, N., Laheru, D., Lau, B., He, J., Zheng, L., Narang, A., Roberts, N.J., Canto, M.I., Lennon, A.M., Goggins, M.G., et al.: Risk of pancreatic cancer in the long-term prospective follow-up of familial pancreatic cancer kindreds. JNCI: Journal of the National Cancer Institute 114(12), 1681–1688 (2022)

25. Rieke, N., Hancox, J., Li, W., Milletari, F., Roth, H.R., Albarqouni, S., Bakas, S., Galtier, M.N., Landman, B.A., Maier-Hein, K., et al.: The future of digital health with federated learning. NPJ digital medicine 3(1), 1–7 (2020)

26. Siegel, R.L., Miller, K.D., Fuchs, H.E., Jemal, A.: Cancer statistics, 2022. CA: A Cancer Journal for Clinicians 72(1), 7–33 (2022), https://doi.org/https://doi.org/10.3322/caac.21708

27. Topaloglu, U., Palchuk, M.B.: Using a federated network of real-world data to optimize clinical trials operations. JCO clinical cancer informatics 2, 1–10 (2018)

28. Videha Sharma, I.A., van der Veer, S., Martin, G., Ainsworth, J., Augustine, T.: Adoption of clinical risk prediction tools is limited by a lack of integration with electronic health records. BMJ Health & Care Informatics 28(1) (2021)

